# Random-walk, agent-level pandemic simulation (RAW-ALPS) for analyzing effects of different lockdown measures

**DOI:** 10.1101/2020.04.29.20084699

**Authors:** Anuj Srivastava

## Abstract

This paper develops an agent-level stochastic simulation model, termed *RAW-ALPS*, for simulating the spread of an epidemic in a confined community. The mechanism of transmission is agent-to-agent contact, using parameters reported for COVID-19 pandemic. When unconstrained, the agents follow independent random walks and catch infections via physical proximity with infected agents. The main goal of the RAW-ALPS simulation is to help quantify effects of preventive measures – timing and durations of lockdowns – on infections, fatalities and recoveries. Three types of lockdown measures are considered: (1) whole population (except essential workers), (2) only the infected agents, and (3) only the symptomatic agents. An infected agent under quarantine can only infect a co-inhabitant, thus causing a decline in infections during lockdowns. The model provides quantifications of changes in infection rates and casualties by imposition and maintenance of restrictive measures in place. The results show that the most effective use of lockdown measures is when all infected agents, including both symptomatic and asymptomatic, are quarantined, while allowing for free movements of all uninfected agents. This calls for regular and extensive testing of the population to isolate and restrict all infected agents.

## 1 INTRODUCTION

There is a great interest in stochastic modeling and analysis of medical, economical and epidemiological data resulting from current the COVID-19 pandemic [2]. From an epidemiological perspective, until a large amount of infection, containment, and recovery data from this pandemic becomes available, the community will rely *primarily on simulation models* to help assess situations and to evaluate countermeasures ([1]). Naturally, simulation systems that follow precise mathematical and statistical models are playing an important role in understanding this dynamic and complex situation ([8]). There have been a large number of models proposed in the past literature, relating to the the spread of epidemics through human contacts or otherwise. They can be broadly categorized in two main classes (a more detailed taxonomy can be found in [15]):

1. **Population-Level Coarse Deterministic Modeling**: A large number of epidemiological models have focused on coarse, population-level summaries - counts of infected (I), susceptible (S), removed or recovered (R), etc. The most popular model of this type is the Susceptible-Infected-Removed (SIR) model [17] proposed by Kermack and McKendrick in 1927. This model uses ordinary differential equations to model constrained growth of the counts in these three categories. There are several other variations of this model including the SIRD model [20] given by:

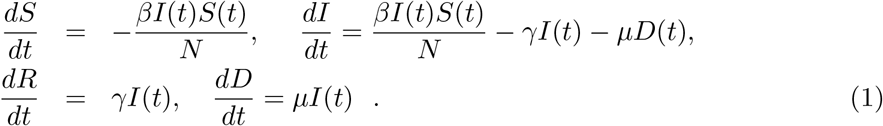

The scalar parameters *β, γ*, and *µ* together control the dynamics of infections, recovery, and mortality. The zero-sum condition 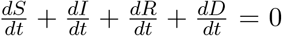 ensures constancy of the community size: *S*(*t*)+*I*(*t*)+*R*(*t*)+*D*(*t*) = *N*. A large number of other papers have studied variants of this basic model and have adapted them for different epidemics, such as ebola and SARS [12]. While there are spatial versions of SIR models, they are not focused on detailed modeling of spatial dynamics. They typically use a uniform static grid to represent the spread of infections, from a location to its neighbors, over time. In general, these models do not explicitly model agent dynamics as residents move around in a community or across communities. Additionally, these models provide deterministic outcomes, with no mechanism to incorporate randomness in the model. Several recent simulation models, focusing directly on COVID-19 illness, also rely on such coarser community level models [22].
2. **Agent-Level Modeling**: While population-level dynamical evolutions of population variables are simple and effective for overall assessment, they do not take into account any social dynamics, human behavior, societal restrictions, and complexities of human interactions explicitly. The models that study these human-level factors and variables, while tracking disease at an individual level, are called *agent-level models* [13]. Here one models the mobility, health status, and interactions of individual subjects (agents) in order to construct an overall population-level picture in a bottom-up way. The advantages of agent-based models are that they are more detailed and one can vary the parameters of restriction measures, such as social distancing, at a granular level to infer overall outcomes. Furthermore, these models have built-in stochastic components for agent motions, interactions, infections, and recoveries, thus enabling a more realistic simulation environment. Agent-based models have been discussed in several papers, including [11, 21, 16, 15] and so on. The importance of simulations-based analysis of epidemic spread is emphasized in [19] but with a focus on infection models within host. Some aspect of spreading of diseases using network contact is also discussed. Hunter et al. [16] construct a detailed agent-based model for spread of infectious diseases, taking into account population demographics and other social conditions, but they do not consider countermeasures such as lockdowns in their simulations. A broad organization of different agent-based simulation methods have been presented in [15]. Although there are numerous other papers on the topic of agent-based simulations for simulating spread of infections, we have only listed the most relevant ones.

In this paper we develop a mathematical simulation model, named *RAW-ALPS*, to replicate the spread of an infectious disease, such as COVID-19, in a confined community and to study the influence of some external interventions on final outcomes. Since RAW-ALPS is purely a simulation model, the underlying assumptions and choices of statistical distributions for random quantities become critical in its success. On one hand it is important to capture the intricacies the observed phenomena as closely as possible, using sophisticated modeling tools. On the other hand, it is important to keep the model efficient and tractable (for individual laptops) by using simplifying assumptions. One can, of course, relax these assumptions and obtain more and more realistic models as desired, albeit with increased computational complexity.

### Simulation Setup

We assume a closed community with infection initiated by a single infected agent at the beginning. The infections are transmitted through physical exposure (proximity) of mobile susceptible agents to mobile infected agents, as shown in the middle panel of Fig.1. When unconstrained, the agents follow a smooth random-walk model independent of other agents (see left panel of Fig.1). When instructed to lockdown, an agent moves towards his/her household unit and stays put until the restrictions are imposed. The households are arranged, equally spaced at the grid points of a square-domain (Fig.1 right)

An agent’s health situation follows the chart shown in the left side of Fig. 2. Susceptible agents become infected by exposure to infected agents with certain probability. The infected agents go through a period of sickness, with two eventual outcomes – full recovery for most and death for a small fraction. That is, one starts as non-infected or susceptible, potentially gets infected with a certain probability, and later recovers or dies according to their event probabilities. Those with non-fatal infections are further labeled as symptomatic or asymptomatic agents. Naturally, agents with fatal outcomes are also labeled as symptomatic. This labeling allows for selective imposition of lockdown measures on either full population or just a subset. Once recovered, we assume that the agents can no longer be infected, as suggested by CDC FAQ [4]. The social dynamical model used here is based on a fixed domicile, i.e. each agent has a fixed housing unit. Under unrestricted conditions, or no lockdown, the agents are free to move over the full domain using a simple motion model. These motions are independent across agents and encourage smooth paths. Under lockdown conditions, the required agents head directly to their housing units and generally stay there during that period. The remaining agents, including a small fraction of agents termed *essential workers*, are allowed to move freely under the restrictions. The infected agents under quarantine can only infect susceptible agents living in the same household and not the general public, as shown in the right side of Fig. 2. Similarly, mobile infected agents can only infect other mobile agents but not those under quarantine.

**Figure 1:**
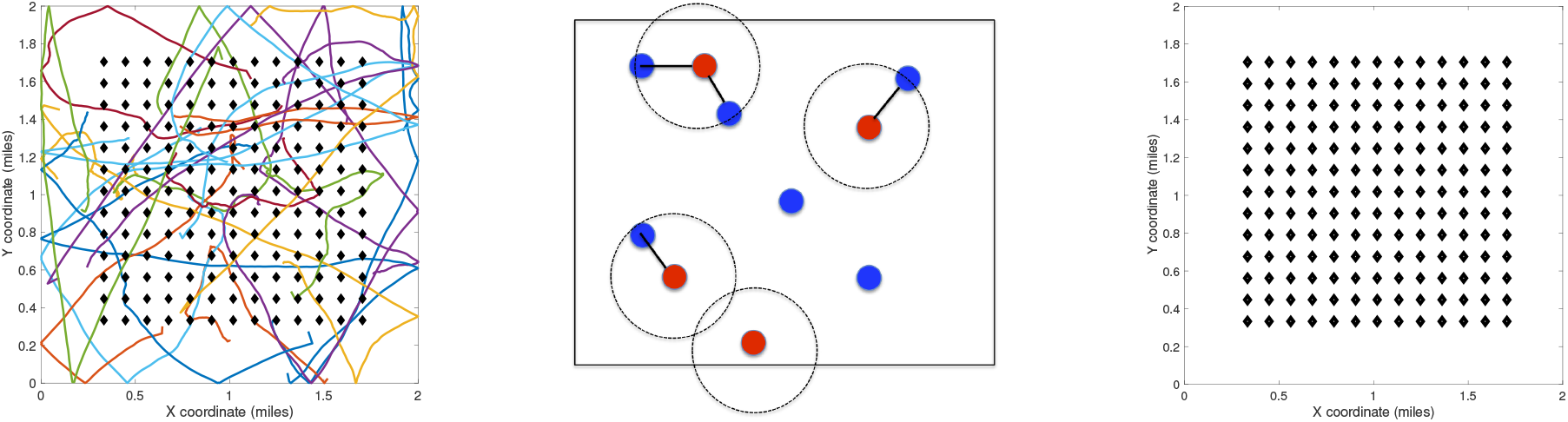
Left: Random walk model for agent motions. Middle: Proximity-based spread of infections from infected to susceptible agents. Right: Layout of households on a square grid in the center of the domain.

**Figure 2:**
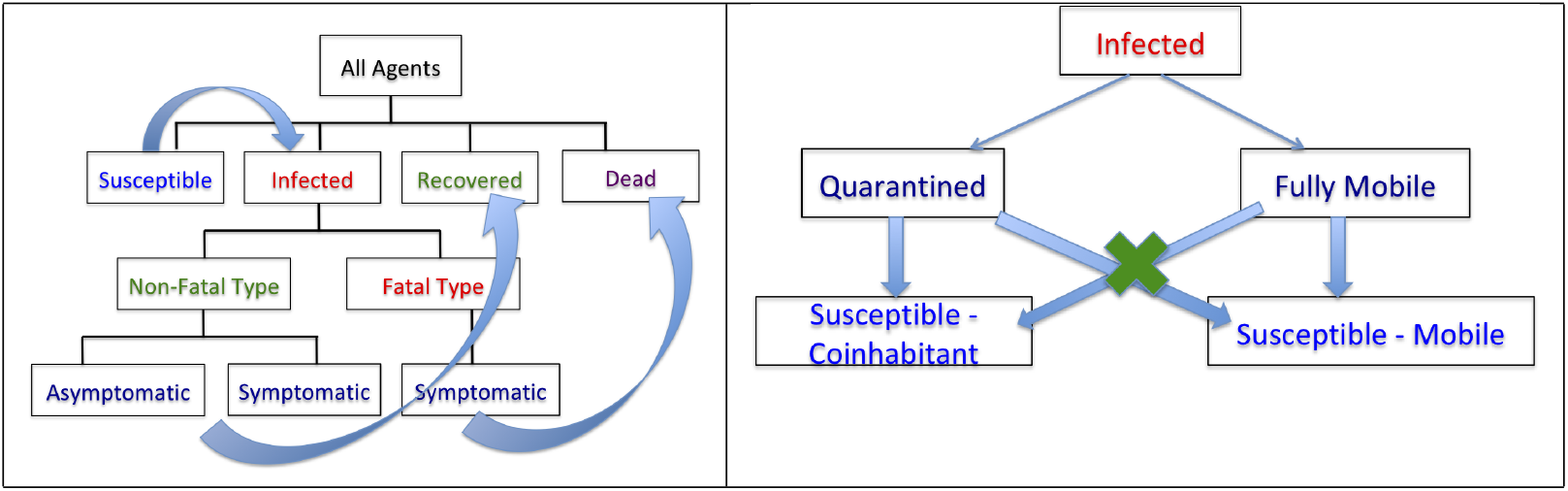
Left: Flow chart for evolution of infection dynamics for the population. Right: Scheme for infection of susceptible agents from infected agents.

The main highlights and contributions of this paper are as follows: (1) Agent-level transmission of infections and thus more granular analysis than (population level) deterministic dynamical models; (2) Random-walk motions of agents when unconstrained and constrained to households under lockdowns; (3) Different classifications of infections – fatal and non-fatal with the latter being either symptomatic or asymptomatic; (4) Selective lockdowns for different strata of population; and (5) Statistical quantifications of gains resulting from lockdown restrictions and their timings on infection rates.

The rest of this paper is organized as follows. Section 2 develops the proposed RAW-ALPS model, specifying the underlying assumptions and motivating model choices. It also discusses choices of model parameters and provides comparisons with the SIRD model. Section 3 presents some illustrative examples and discussed computational complexity of RAW-ALPS. The use of RAW-ALPS in understanding influences of countermeasures is presented in Section 4. The paper ends by discussing model limitations and suggesting some future directions.

## 2 Random-Walk, Agent-Level Pandemic Simulation (RAW-ALPS)

In this section we develop our simulation model for agent-level interactions and spread of the infections across a population in a well-defined geographical domain. In terms of the model design, there are competing requirements for such a simulation to be useful. On one hand, we want to capture detailed properties of agents and their pertinent environments so as to render a realistic model of pandemic evolution with or without countermeasures. On the other hand, we want to keep model complexity reasonably low, in order to utilize it for analyzing variable conditions and countermeasures. Also, to obtain statistical summaries of pandemic conditions under different scenarios, we want to run a large number of simulations and compute averages. This also requires keeping the overall model simple, from a computational perspective, to allow for multiple runs of RAW-ALPS.

### 2.1 Simplifying Assumptions

The overall setting of the simulation model is as follows. We assume that the community is located in a square geographical region *D* with *h* household units arranged centrally on a uniformly-spaced square grid in *D*. We assume that there is a fixed number *N* of total agents in the community (including all classifications – uninfected, infected, dead, etc) and their health status updates every unit interval (one hour) indexed by variable *t*. The agents freely traverse all parts of *D* when unconstrained but are largely restricted to their home units under restrictions. Next, we specify some simplifying assumptions:

- **Independent Agents**: Each agent (person) has independent motion and independent infection status. The actual infection event is of course dependent on being in a close proximity of an infected carrier (within a certain distance, say *≈* 6 feet) for a certain exposure time. But the probability of an agent being infected is independent of such events for other persons.
- **Full Mobility In Absence of Restrictive Measures**: We assume that each agent is fully mobile and moves across the domain freely when no restrictive measures are imposed. In other words, there is no effect of an agent’s age, gender, or health on his/her mobility. Also, we do not impose any day/night schedules on the motions. Some papers, including [10], provide a two- or three-state models where the agents transition between some stable states (home, workplace, shopping, etc) in a predetermined manner. To implement such detailed models requires careful considerations about the daily movement patterns of agents, and that increases model complexity.
- **Homestay During Restrictive Measures**: We assume that most agents stay at home at all times during the restrictive conditions. Only a small percentage (set as a parameter *ρ*_0_) of population, representing essential workers, are allowed to move freely but the large majority stays at home.
- **Sealed Region Boundaries**: In order to avoid complications resulting from a transportation model in the system, we assume that there is no transfer of agents into and out of the region *D*. The region is modeled to have reflecting boundaries to ensure that all the citizens stay with in the region. The only way the population of *D* is changed is through death.
- **Fixed Domicile**: The whole community is divided into a certain number of living units (households/buildings). These units are placed in square blocks with uniform spacing. Each agent has a fixed domicile at one of the units. During a lockdown period, the agents proceed to and stay at home with a certain fidelity. We assume that all agents within a unit are exposed to each other, i.e. they are in close proximity, and can potentially infect others.
- **No Re-Infection**: We assume that once a person has recovered from the diseased then he/she can not be infected again for the remaining observation period. While this is an important unresolved issue for the current COVID-19 infections, it has been a valid assumption for the past Corona virus infections and it remains the current CDC guideline [4].
- **Single Patient Ground Zero**: The infection is introduced in the population using a **single carrier**, termed patient ground zero at time *t* = 0. This patient is selected randomly from the population and the time *t* is measured relative to this introduction event.
- **Constant Immunity Level**: The probability of infection of agents, under the exposure conditions, remains same over time. We do not assume any increase or decrease in agent immunity over time. Also, we do not assign any age or ethnicity to the agents and all agents are assumed to have equal immunity levels.

### 2.2 Model Specifications

There are several parts of the model that require individual specifications. These parts include a model on dynamics of individual agents (with or without restrictions in place), the mechanisms of transmitting infections from agent to agents, and the process of recovery and fatality for infected agents. A full listing of the model parameters and some typical values are given in Table 2 in the appendix.

- **Motion Model**: The movement of an agent follows a simple model where the instantaneous velocity *v*_*i*_(*t*) is a weighted sum of three components: (1) velocity at the previous time, *i*.*e. v*_*i*_(*t −* 1), (2) a directed component guiding them to their home, (*h*_*i*_ *− x*_*i*_(*t −* 1)), where *x*_*i*_(*t*) is the agent location at time *t*, and (3) an independent Gaussian increment *σw*_*i*_(*t*), *w*_*i*_(*t*) *∼ N* (0, 1). Note that the motions of different agents are kept independent of each other. The location *h*_*i*_ *∈ D* denotes the home unit (or stable state) of the *i*^*th*^ person. Using mathematical notation, the model for instantaneous position *x*_*i*_(*t*) and velocity *v*_*i*_(*t*) of the *i*^*th*^ agent are given by:

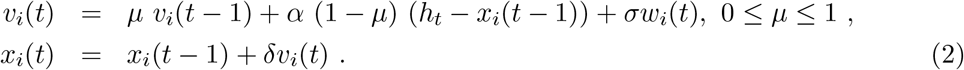

Here *α ∈* R_+_ determines how fast one moves towards their home, and *µ* quantifies the degree to which one follows the directive to stay home. If *µ* = 0, then the person reaches home and stays there, except for the random component *w*_*i*_. However, if *µ* = 0.5, then a significant fraction of motion represents continuity in velocity, irrespective of the home location. The value *µ* = 1 implies that either there is no lockdown or the person does not follow the directive. **Reflecting Boundaries:** When a subject reaches boundary of the domain *D*, the motion is reflected and the motion continues in the opposite direction. Fig. 3 shows examples of random agent motions under different simulation conditions. The leftmost case shows when there is no lockdown and the agents are moving freely throughout. The middle case shows the case when the restrictions are imposed on Day 10 and the restrictions stay in place after that. The last plot shows the case where a lockdown is imposed on Day 10 and then lifted on Day 20. The blue curves represent free motion before a lockdown, red denotes motion towards home during a lockdown, and green represents free motion again after the lockdown is lifted.

**Figure 3:**
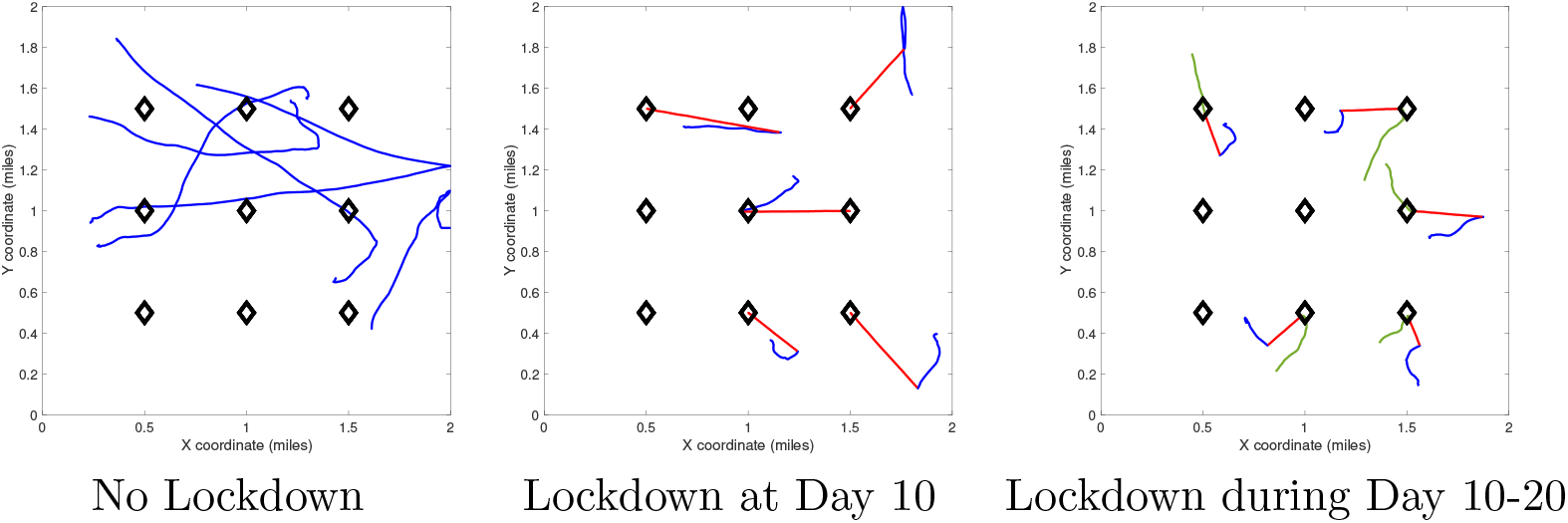
Sample agent motions under different conditions. Blue curves denote unrestricted movements, red curves denote movement during lockdown, and green curves denote movement after lockdown.
- **Restrictions (or Lockdown) Model**: Once the lockdown starts, at time *T*_0_, agents are directed towards their homes and asked to stay there. One can impose restriction on the whole population or a certain subset (say symptomatic agents). We assume that *ρ*% of the subject follow this directive while the others ((100 *− ρ*)%) follow a different motion model. The variable *ρ* changes over time according to: 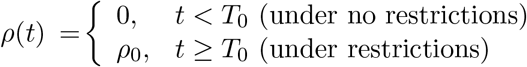. We note that the people who do not follow restrictions follow the prescribed motion model (Eqn. 2) with *µ* = 1.
- **Exposure-Infection Model**: Infection of an agent depends on the level of exposure to another infected person. Recall that mobile infected agents can only infect other mobile agents while quarantined agents can only infect agents in the same household. Furthermore, this process is controlled by the following parameters:
  - The separation between the subject and the infected person is less than *r*_0_.
  - The amount of exposure in terms of the number of time units is at least *τ*_0_. We use the cumulative time exposure over the whole history, rather than just the recent history.
  - Under these conditions, the probability of being infected at each time *t* is an independent Bernoulli random variable with probability *p*_*I*_.
- **Recovery-Death Model**: Once a subject is infected, we randomly assign a label immediately. Either an infected agent is going to recover (non-fatal type, or NFT) or the person is eventually going to die (fatal type, or FT). The probability of having a fatal infection given that a person is infection is *p*_*F*_. An agent with NFT label is further classified as either Symptomatic (S) or Asymptomatic (AS) with a certain probability *p*_*S*_. All agents with FT label are classified as symptomatic.
  - **Recovery**: A subject with a non-fatal type (NFT) is sick at least for a period of *T*_*R*_ days. After this period, the person can recover at any time according a Bernoulli random variable with probability *p*_*R*_.
  - **Fatality**: A subject with a fatal type (FT) is sick for a period of *T*_*D*_ days. After this period, the person can die at any time independently according a Bernoulli random variable with probability *p*_*D*_.

### 2.3 Chosen Parameter Values

A complete listing of the ALPS model parameters is provided in the Appendix in Table 2. In this section, we motivate the values chosen for those parameters in these simulations. These values are motivated by the current reports of COVID-19 pandemic. As argued in [14], a realistic choice of parameters is very important in establishing validity of simulation models.

- **Population Density**: We have used a square domain of dimensions 2 miles *×* 2 miles for a community with population of N agents. The value of *N* changes in different experiments. For *N* = 900, the model represents a population density of 225 people/mile^2^. The community contains *h* living units (buildings) with a domicile of *N/h* people per unit. In case *N/h* is high (*≈* 100), a unit represents a tall building in metropolitan areas, but when *N/h* is small (*≈* 5), a unit represents a single family home in a suburban area. The time unit for updating configurations is one hour and occurrence of major events is specified in days. For example, the lockdown can start on Day 1 and end on Day 30.
- **Agent Speeds**: The standard deviation for accelerations in agent mobility are approximately 1-5 feet/hour (fph). Through integration over time, this results in agent speeds up to 1000 fph. We assume that *ρ*_0_ = 0.9 *−* 0.98, *i*.*e*. 90-98% of the people follow the restriction directives.
- **Infection Rates**: The physical distance between agents to catch infection should at most *r*_0_ *≈* 6*ft* and the exposure time should be at least *τ*_0_ = 5 time units (hours). The probability *p*_*I*_ of getting infected, under the right exposure conditions, is set at 5% at each time unit (hour) independently. There is no current reference literature on selecting this value, since it is difficult to measure precise exposure events for people who have tested positive for COVID-19. While contact tracing [5] is being developed to ascertain infection rates, there is no public data currently to measure this infection rate. This value leads to overall infection rates that are similar to national and international infection rates [9].
- **Fatality Rate**: Once infected, the probability of having a fatal outcome is set at 5-10%, according to the mortality rate listed by CDC [6]. The time period of recovery for agents with non-fatal outcomes starts at 7 days. The probability of reaching full recovery for those agents is *p*_*R*_ = 0.001 at each subsequence time unit (hour). Similarly, for the agents with fatal outcomes, the period of being infected is set to be 7 days and after that the probability of death at each time unit (hour) is set to be *p*_*D*_ = 0.1.
- **Asymptomatic Infected Agents**: The percentage of infected agents who remain asymptomatic is set to be in the range 15 *−* 40% [3, 18].

Figure 1 provides a pseudocode for the RAW-ALPS algorithm.

**Given**: Simulation Parameters: Interval length *T*, Population size *N*, and other model parameters as discussed in paper; Lockdown Periods *T*_0_ *− T*_1_; Lockdown type: Full, Infected, or Symptomatic Only

**Result**: Locations and infection status of each agent over the observation interval; Cumulative rate curves for S, I, R, and D.

Initialize the locations *x*_*i*_ ∈ *D* uniformly. Select an index *i*_0_ *∈* {1, 2, …, *N*} randomly to be the seed infected agent;

Initialize the lists *L*_*I*_ = {*i*_*o*_}, *L*_*A*_ = {1, …, *N*}, *L*_*S*_ = *L*_*A*_*/L*_*I*_, *L*_*D*_ = [], *L*_*R*_ = [];

#### Algorithm 1 RAW-ALPS Pseudocode.

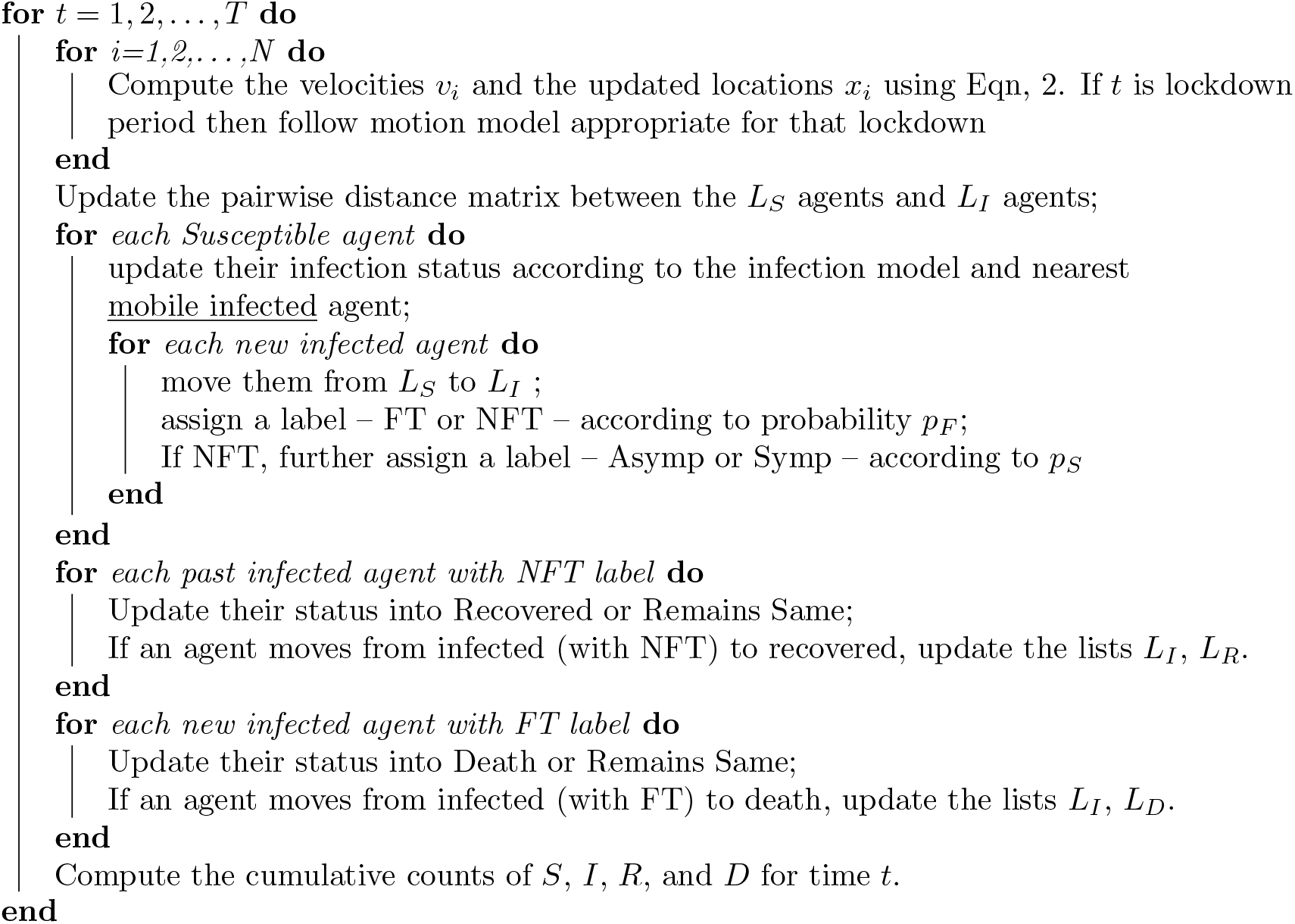

### 2.4 Model Validation

Although RAW-ALPS is perhaps too simple model to capture intricate dynamics of an actual active society, it does provide an efficient tool for analyzing effects of countermeasures during the spread of a pandemic. Before it can be used in practice, there is an important need to validate it in some way.

As described in [15], there are several ways to validate a simulation model. One is to use real data (an observed census of infections over time) in a community to estimate model parameters, followed by a statistical model testing. While such data may emerge for COVID-19 for public use in the future (especially with the advent of tracking apps being deployed in many countries), there is currently no such agent-level data available for COVID-19. The other approach for validation is to consider coarse population-level variables and their dynamics, and compare them against established models such as SIR and its variations. We will perform validation of RAW-ALPS in two ways.

#### 2.4.1 Qualitative Comparisons with SIR Model

As the first comparison, we study *shapes* of infection curves resulting from the RAW-ALPS model and compare them qualitatively to the shapes resulting from SIR model. Fig. 4 shows plots of the evolutions of global infection counts (susceptible, infection, recovered) in a community under the well-known SIR model (on the left) and the proposed ALPS model (on the right). In the ALPS model the counts for recovered and fatalities are kept separate, while in SIR model these two categories are combined. One can see a remarkable similarity in the shapes of corresponding curves and this provides a certain validation to the RAW-ALPS model. In fact, given the dynamical models of agent-level mobility and infections, one can derive parameters of the population-level differential equations used in the SIRD model. We pursue this topic in the next section.

**Figure 4:**
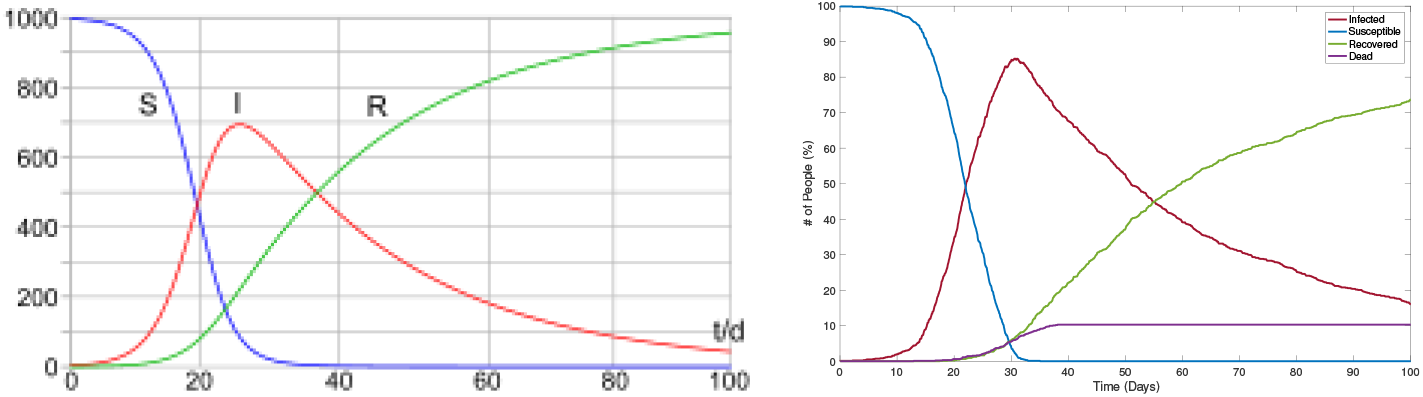
The evolution of population-level infection measurements under a typical SIR model (left), source: wikipedia) and the RAW-ALPS model (right). Visually, RAW-ALPS model is able to generate infection curves with shapes similar to that of SIR model.

### 2.4.2 Estimation of SIRD Parameters

In this section we use data simulated from RAW-ALPS to fit the SIRD model given in Eqn. 1 and use estimated SIRD model parameters to provide interpretations. Re-arranging equations in SIRD model (Eqn. 1), we get:

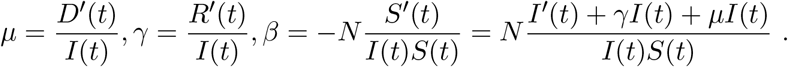

Let 1_*f*(*t*)*>*0_ denote the domain over which a function *f*(*t*) is strictly positive. We define the estimators of SIRD parameters to be:

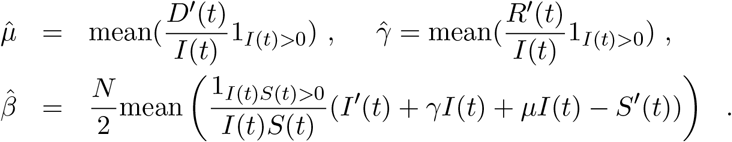

Since in our simulations, we have a small population size (*N*), the number of dead agents becomes a constant soon after any changes are complete, making *D′*(*t*) = 0 for most of the study. In order to focus on mortality rate in the active period, we modify the estimator of *µ* to be: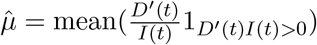.

Fig. 5 shows an example of this estimation using data simulated from RAW-ALPS for an unrestricted situation. The left plot shows the evolution of *S/I/R/D* functions under the chosen simulation parameters. The infection reaches its peak around Day 26 and there are no uninfected agents left after that. For this data, the estimated values of parameters are 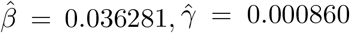, and 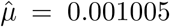 (at hourly unit time). We clarify that the simulation is updated at an hour unit but the final estimates of these rates are converted into daily time unit to compare with published values. The basic reproduction number for this simulation comes out to be 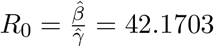

**Figure 5:**
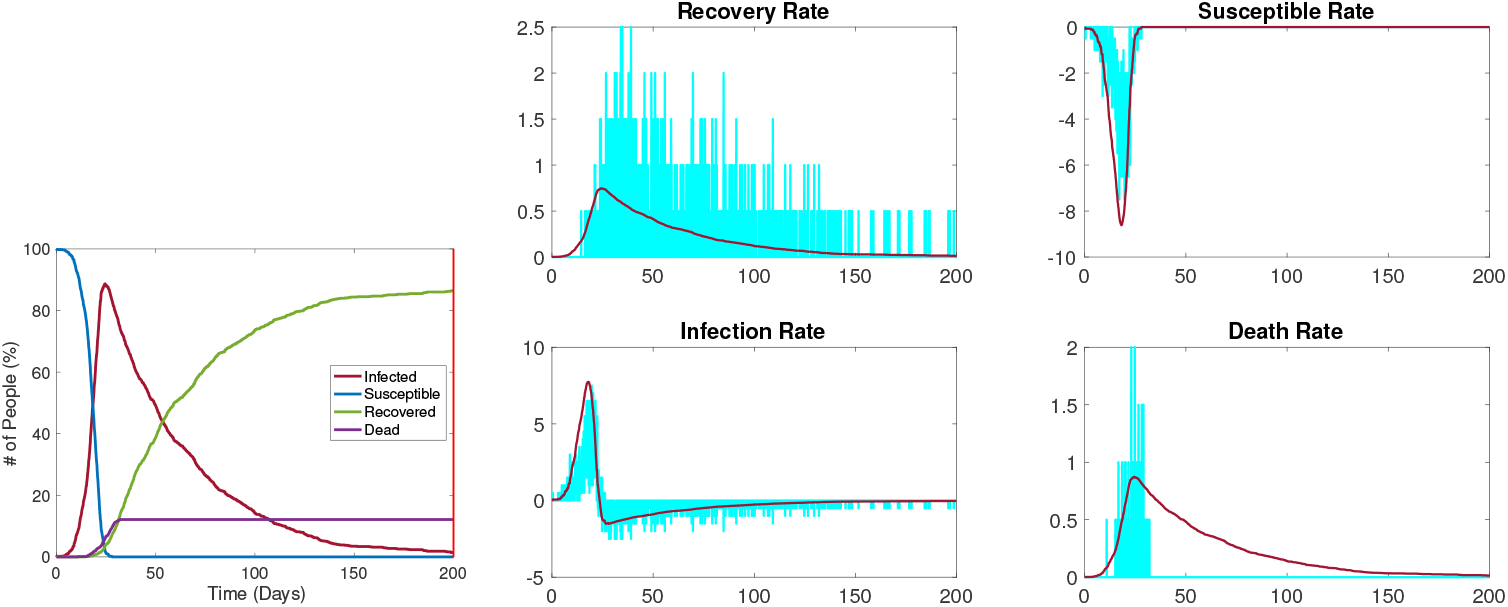
SIRD parameter estimation from data simulated by RAW-ALPS for an unrestricted spread.

Fig. 6 shows when the restrictions are imposed on Day 5. The left plot shows the evolution of *S/I/R/D* functions – The infection reaches its peak around Day 20 even though there is a significant portion of uninfected agents. For this data, the estimated values of parameters are 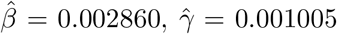, and 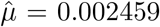 (at hourly unit time). The basic reproduction number for this simulation comes out to be 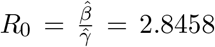. Compared with the previous example we see that the infection rate comes down significantly – from 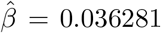 to 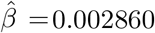 when the restrictions are imposed. Similarly, the reproduction number comes down from 42.1703 to 2.8458.

**Figure 6:**
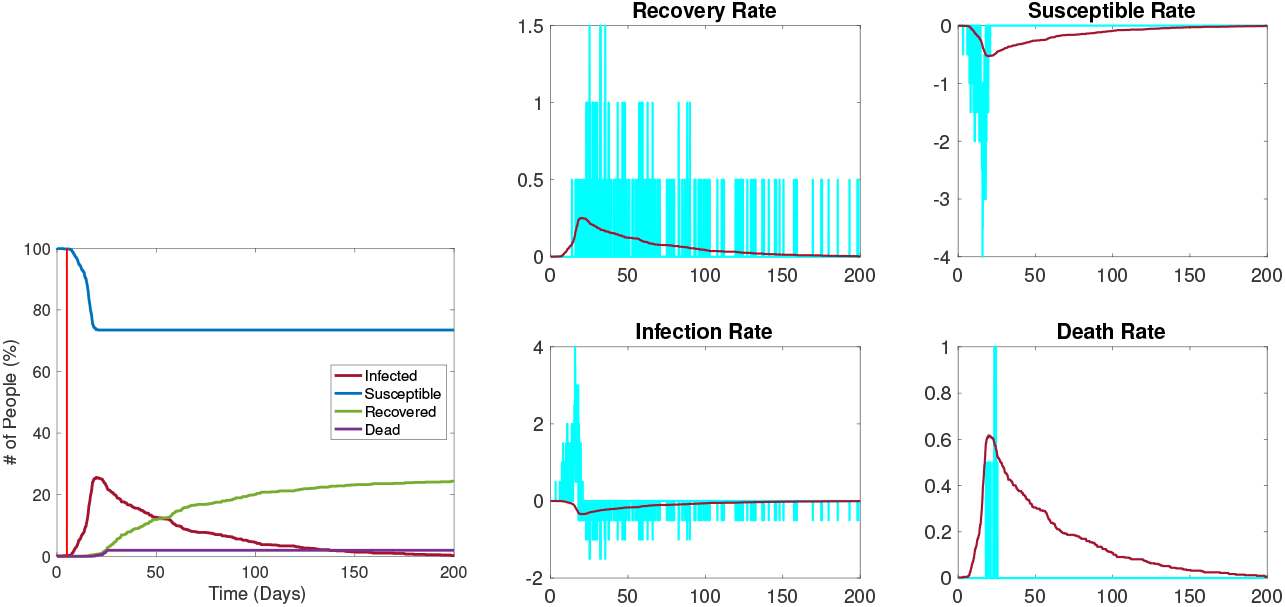
SIRD parameter estimation from data simulated by RAW-ALPS for the case when the restrictions are imposed on Day 5.

## 3 Exemplar Outcomes & Computational Cost

We illustrate the use of RAW-ALPS model by presenting outcomes under some typical scenarios. Further, we discuss the computational cost of running RAW-ALPS on a laptop.

### 3.1 Examples from RAW-ALPS

We start by showing sample outputs of RAW-ALPS under some interesting settings. In these examples, we use a relatively small number of agents (*N* = 1500) and household units (*h* = 289), with *T* = 50 days, in order to improve visibility of displays.

1. **Example 1 – No Restrictive Measures**: Fig. 7 shows a sequence of temporal snapshots representing the community at different times over the observation period. In this example, the population is fully mobile over the observation period and no social distancing restrictions are imposed. The snapshots show the situations at 8, 17, 33, and 42 days. The corresponding time evolution of global count measures is shown in the bottom right panel. The infection starts to spread rapidly around the Day 5 and reaches a peak infection level of 78% around Day 17. Then the recovery starts and continues until very few infected people are left. In this simulation, the number of fatalities is found to be 6%.

**Figure 7:**
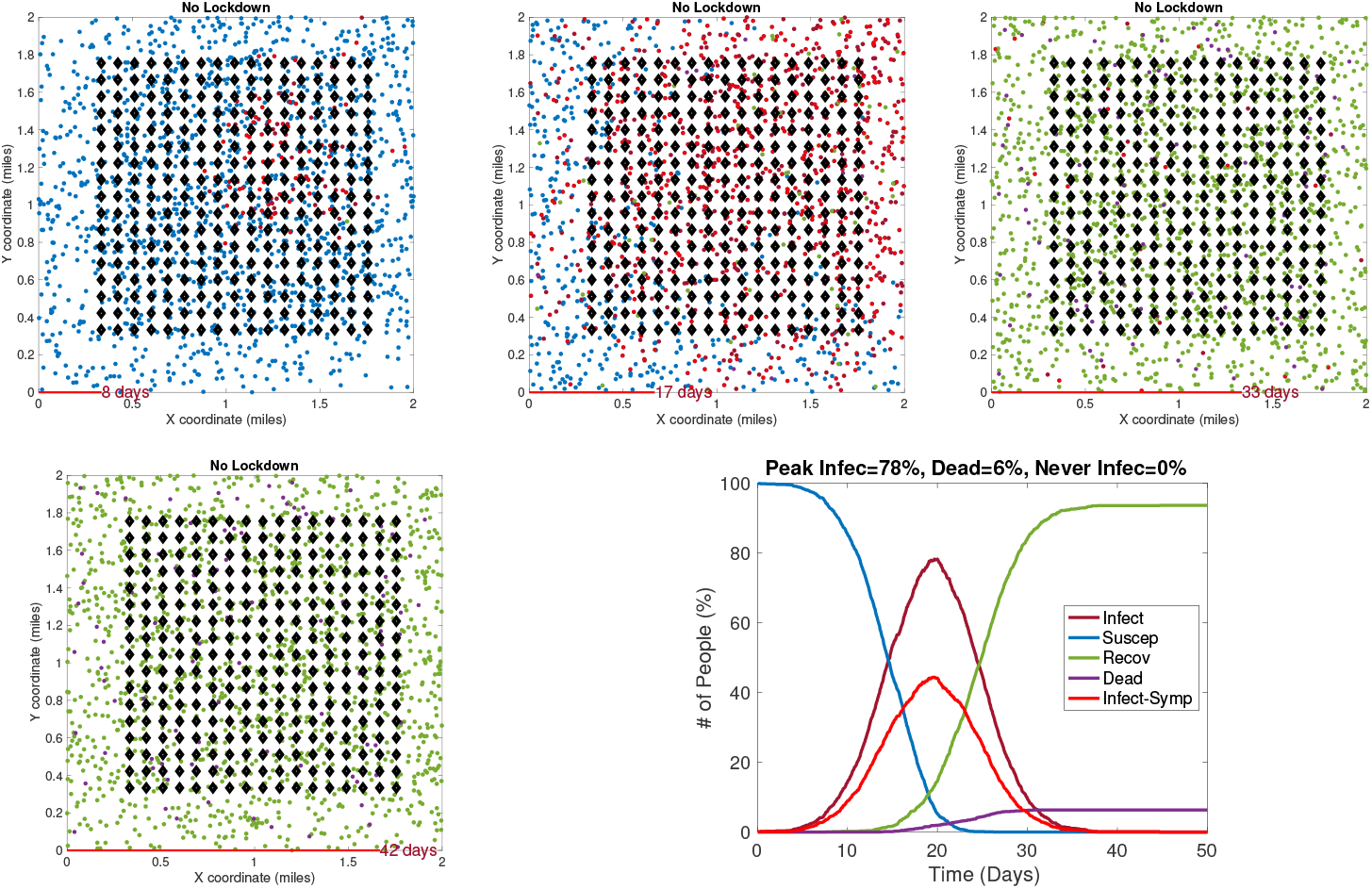
Example 1: Model outputs at different times under no restrictive measures. Blue dots are susceptible agents, red dots are infected agents, green dots are recovered agents, and purple circles denote fatalities.
2. **Example 2 – Early Restrictions**: In the second example, a lockdown of infected agents is introduced on Day 5 and these measures stay in place after that. The results are shown in Fig. 8. Once again we show snapshots for situations at 8, 17, 33, and 42 days. The corresponding time evolution of global count measures is shown in the bottom right panel. As the summary shows, an early restriction of lockdown is very effective in controlling the spread of infection and the peak infection rate is miniscule at 3%.

**Figure 8:**
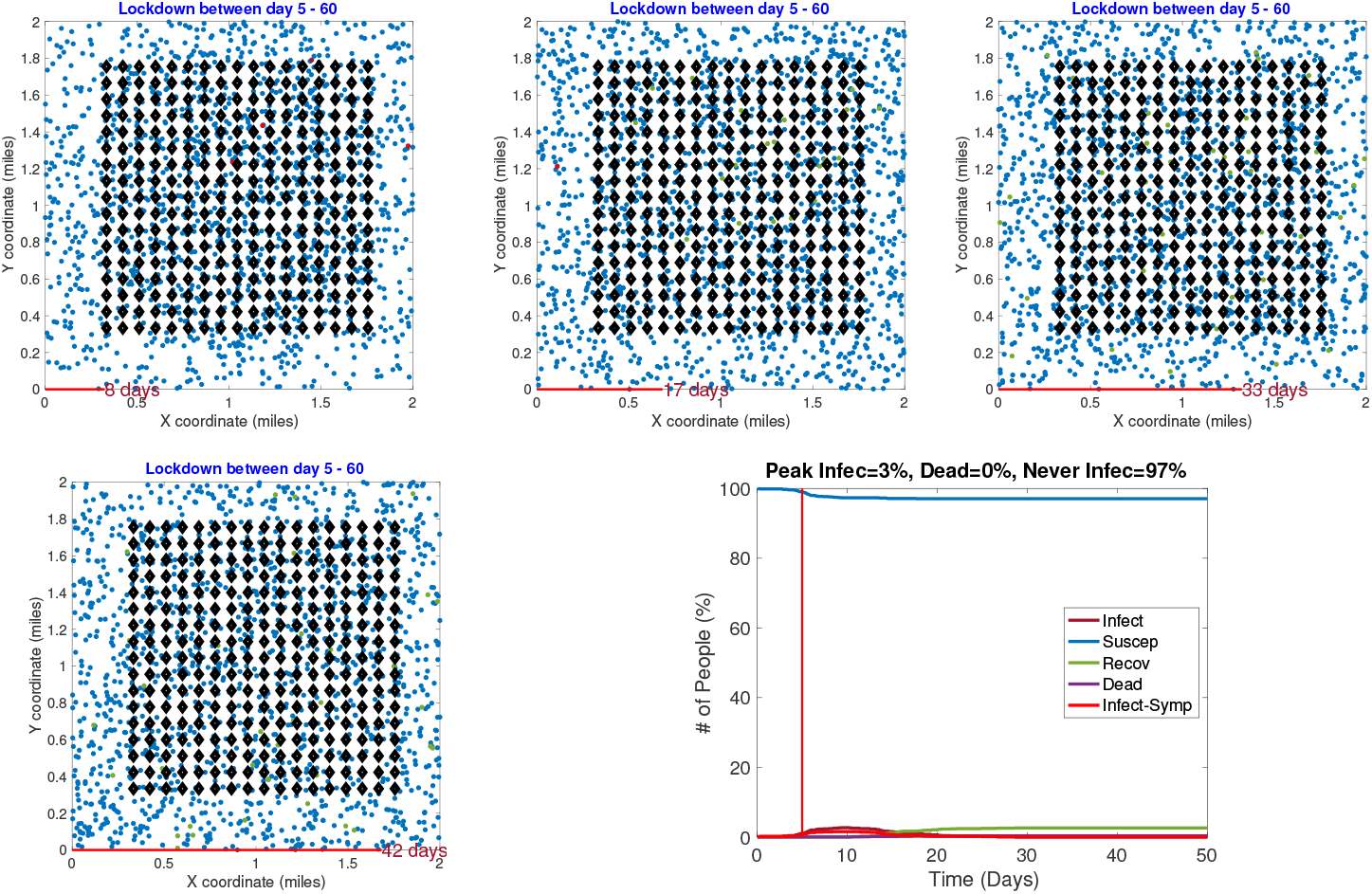
Example 2: Model outputs when a lockdown of infected people is imposed very early (on Day 5).
3. **Example 3 – Early Restrictions but Removed Later**: In the next example, a full lockdown is introduced on Day 5 but removed on Day 30. The results are shown in Fig. 9. In early snapshots, the population is under a lockdown and the spread is minimal. By Day 33, the lockdown is released and the population is fully mobile. Consequently, the infection begins to spread and by Day 42 the infection reaches its peak value.

**Figure 9:**
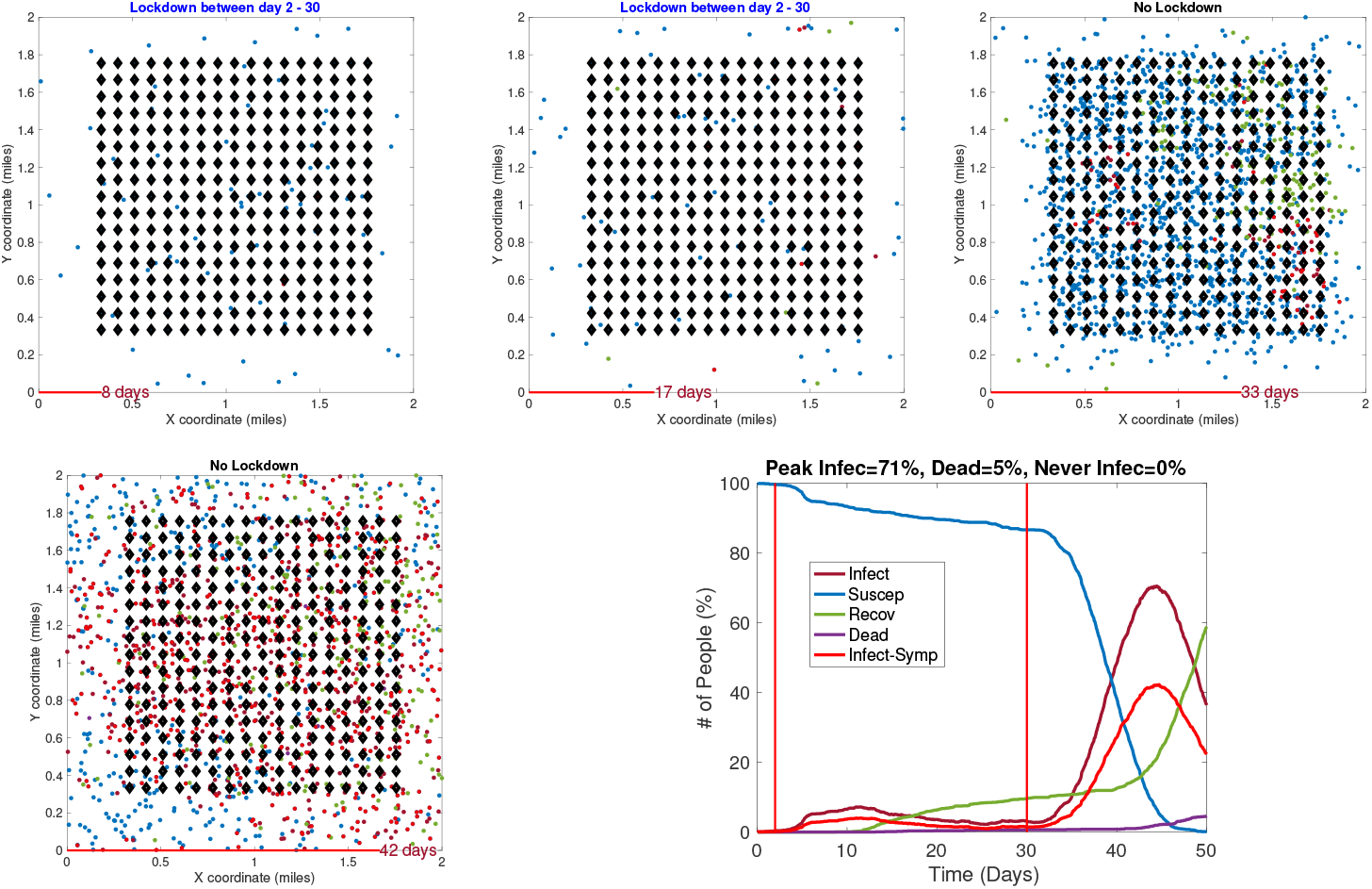
Example 3: Model outputs when a full lockdown is imposed on Day 2 and removed on Day 30.
4. **Example 4 – Later Restrictions** : In the last example, with results shown in Fig. 10, a full lockdown is introduced very late (on Day 10). In this simulation run, the infection rate reaches a peak value of 80% despite a lockdown. This is because the infection had already spread extensively in the population by the time the lockdown starts.

**Figure 10:**
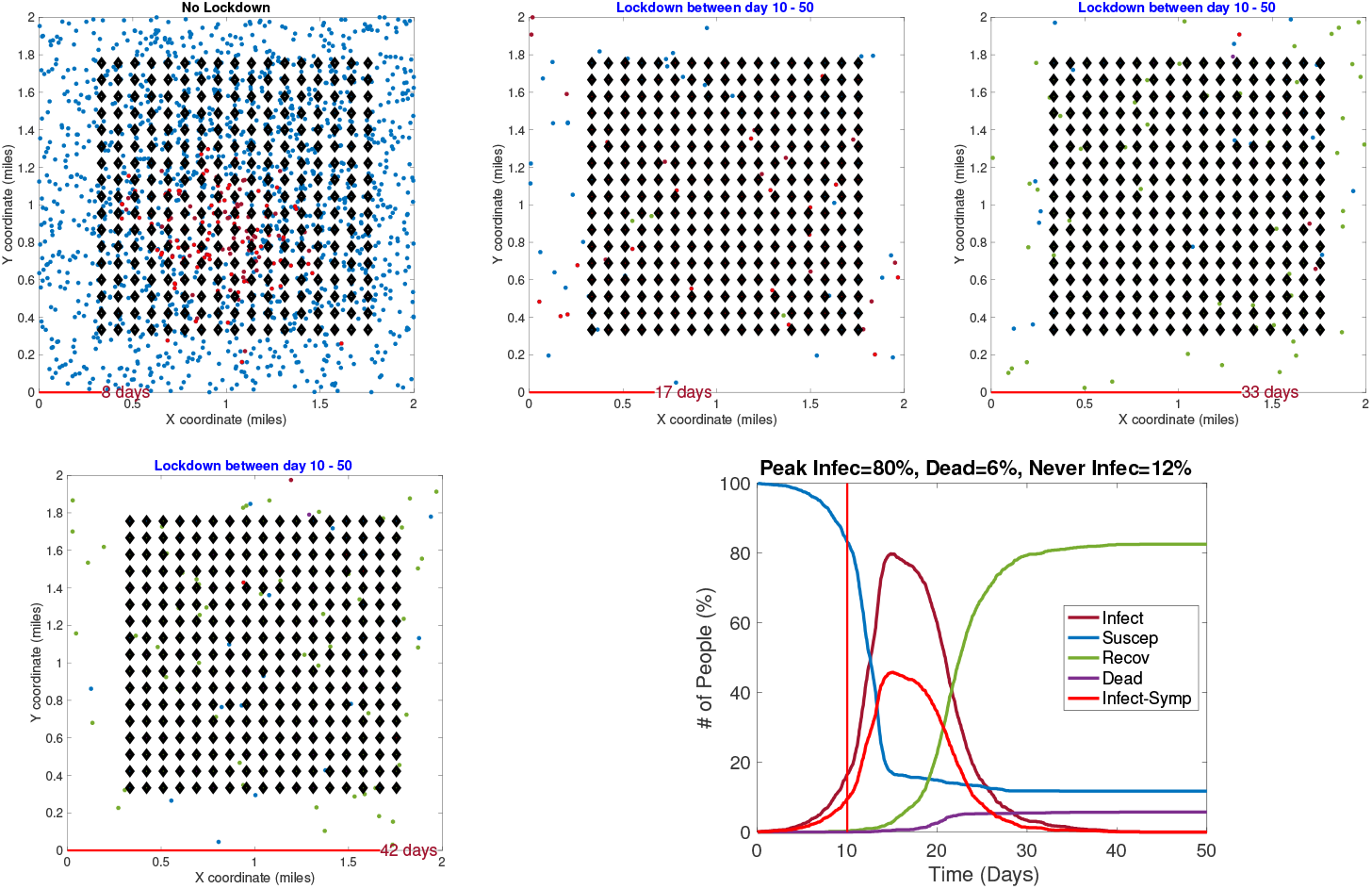
Example 4: Model outputs when a full lockdown is imposed on Day 10.

### 3.2 Computational Complexity

The main computational cost in the simulation comes from the need to update the following variables at each time *t*:

- Locations of each model according to their independent motion model;
- Pairwise distance matrix between Susceptible agents and Infected agents; and,
- Infection status of each agent according to the infection dynamics.

In general we obtain some efficiency by performing matrix operations, rather than using for loops for these updates across agents. Additionally, we increase speed by maintaining a *neighborhood* for each agent and checking interactions only between the neighbors at each *t*.

Since computational efficiency of the simulator is of vital importance, we study the computational cost of running ALPS for different variable sizes. In these experiments, we note the time taken by ALPS code on a McBook Pro laptop with Intel 2.8 GHz Core i7 processor and 16GB memory. In Fig. 11 we plot average run times (using five runs in a setting) of the code for different values of *N, h*, and *T*. Recall that *h* is the number of households units, *N* is the number of agents and *T* is the period length. From these results we are see that the computational cost is linear in *T*, which makes sense. Keeping other variables fixed, the computational cost is super linear in *N*. This is because an increase in *N* represents a higher density and increased infection rates, thus requiring additional computations for tracking diseased agents. Interestingly, for the range of parameter values studies here, the computational cost does not grow with an increase in *h*. As an aside, we note that for *N* = 5000 and *h* = 1521, the run time for *T* = 100 days is approximately 330s and for *T* = 150 days is approx. 484s.

**Figure 11:**
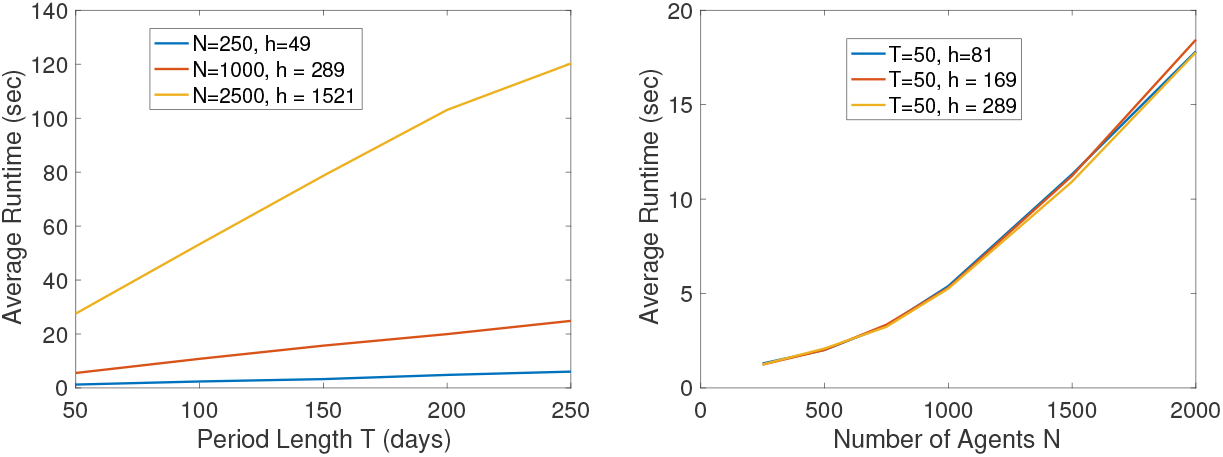
Plots of run times versus simulation parameters *T, N*, and *h*.

## 4 Analyzing Effects of Lockdown Measures

There are several ways to utilize this model for prediction, planning and decision making. We illustrate some of these ideas using examples.

### 4.1 Timing of Imposition of Full Lockdown

First, we study the effect of time of the *full* lockdown on the epidemic infections. In the following simulations, we have used *N* = 1000 agents with *h* = 361 households in the scene domain [0, 2]^2^ miles. For the infection parameters, we use a contact radius *r*_0_ = 0.02 miles, contact period *τ*_0_ = 5 hours and *p*_*I*_ = 0.1. The motion parameters are: *σ* = 0.0002mph, *µ* = 0.02, and *α* = 0.1. In each setting, we run the code 30 times and collection the simulation outputs.

Fig. 12 shows examples of RAW-ALPS outputs when a full lockdown is imposed on the community but at different times. From top to bottom, the plots show lockdowns starting on Day 2, Day 10, and Day 30, respectively, with the last row showing results for no lockdown. Once the restrictions are imposed, they are not removed in these examples. As expected, the best results are obtained for the earliest imposition of restrictions. In the case of no lockdown, the peak infection rate in the population ranges from 60-80% which is a very high number for a community. The fraction of fatalities ranges from 6-8% and the fraction of community that is never infected is zero in all runs. In case the restrictions are imposed on Day 2, with all other parameters held same, there is a remarkable improvement in situation. The peak infection goes down to 2-3%, the fatalities decrease to 0.1-0.2% and the fraction of uninfected goes up to 98-99%. Thus, an early imposition of full lockdown measures helps significantly reduce infection in the community.

**Figure 12:**
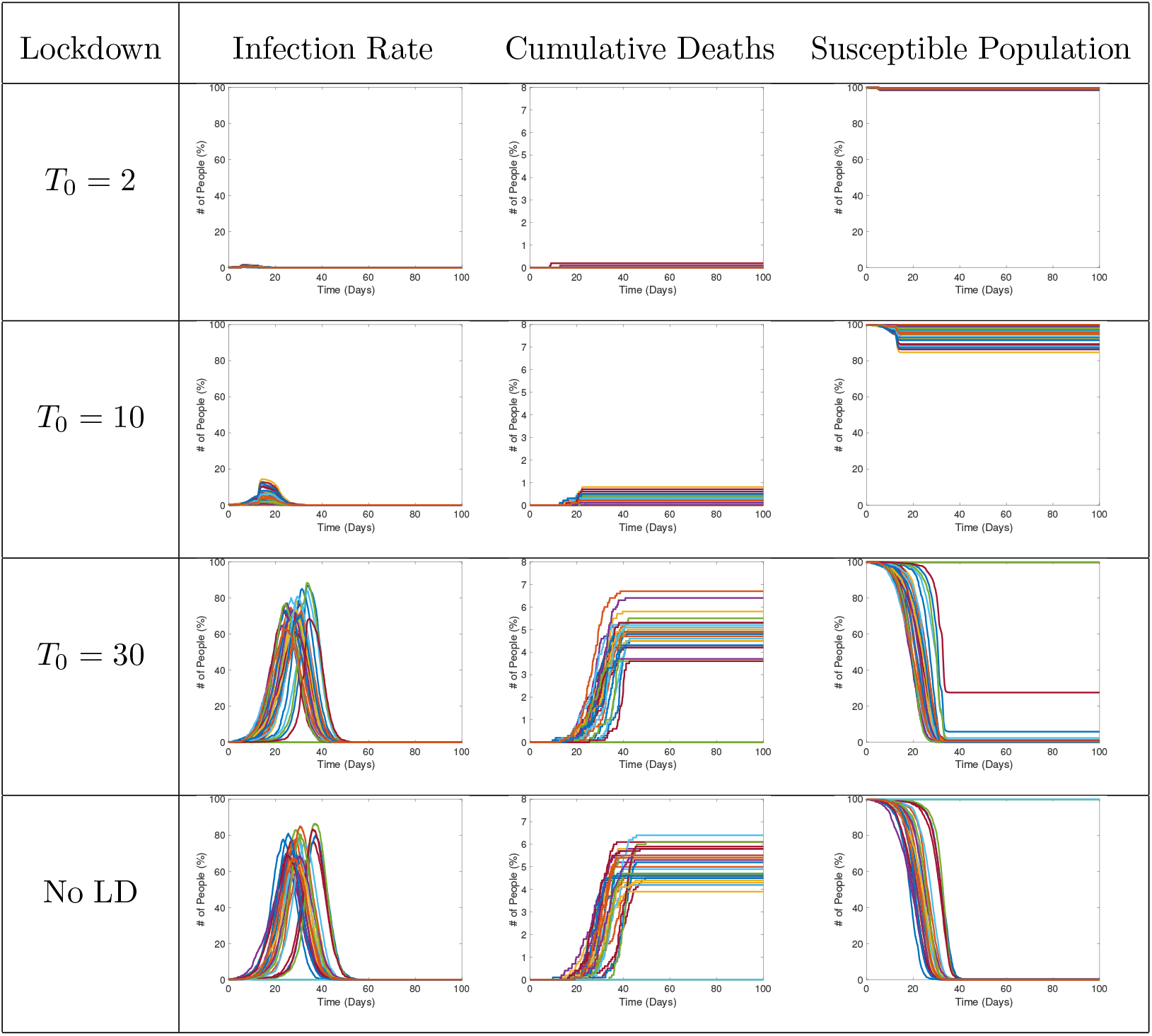
Results from RAW-ALPS runs for a full lockdown starting at different times. From top to bottom, the lockdowns are imposed later and later.

### 4.2 Timing of Removal of Restrictions

In the next set of simulations, we study the effects of lifting restrictions and thus re-allowing full mobility in the community. Some sample results are shown in Fig. 13. Each plot shows the evolution for a different end time *T*_1_, while the start time is kept fixed at *T*_0_ = 5. As these plots indicate, the gains made by early imposition of restrictions are nullified when the restrictions are lifted. In the case of early lifting of restrictions, the full population gets infected eventually. Since we do no assume any change in the immunity levels of the agents over time, the results from early lifting of restrictions are similar to not imposing any restrictions in the first place. The results appear to be same except for being shifted in time.

**Figure 13:**
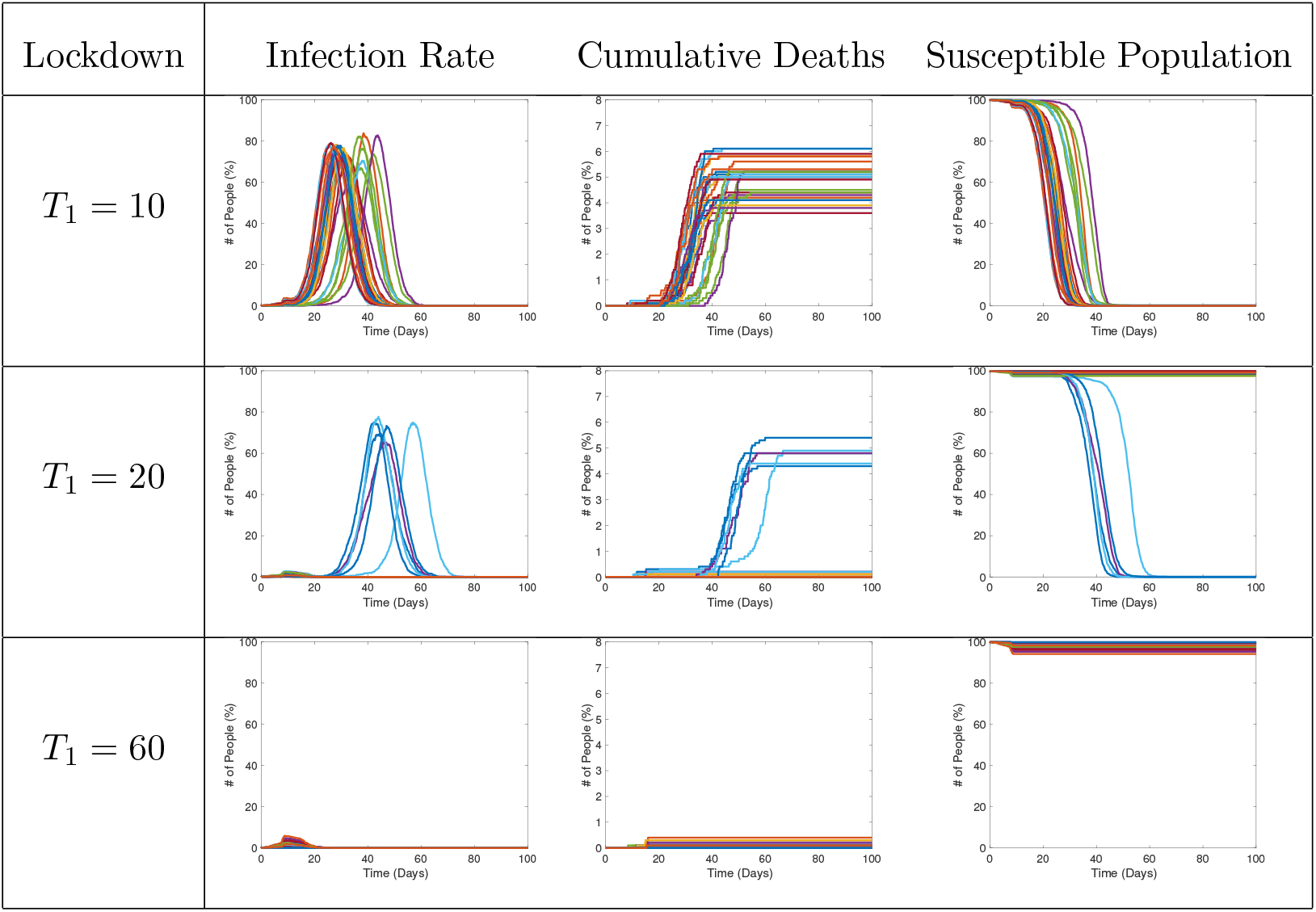
Results from RAW-ALPS runs for a full lockdown starting at different times. From top to bottom, the lockdowns are imposed later and later.

### 4.3 Statistical Summaries

In the next set of experiments, we compute average values of some variables of interest using 30 runs of RAW-ALPS. In the first result, we study three variables – number of deaths, number of people remaining uninfected, and the peak infection rate – using *N* = 1000 agents living in a community of *h* = 361 households, observed over [0, 100] days. We vary the start time *T*_0_ (start day of restrictions) from 2 to 30 and then to 100 and study the resulting outcomes. (The value of *T*_0_ = 100 implies that the restrictions are never imposed in that setting.) Fig. 14 shows box plots of the three varying with *T*_0_ with each row representing different type of lockdown.

**Figure 14:**
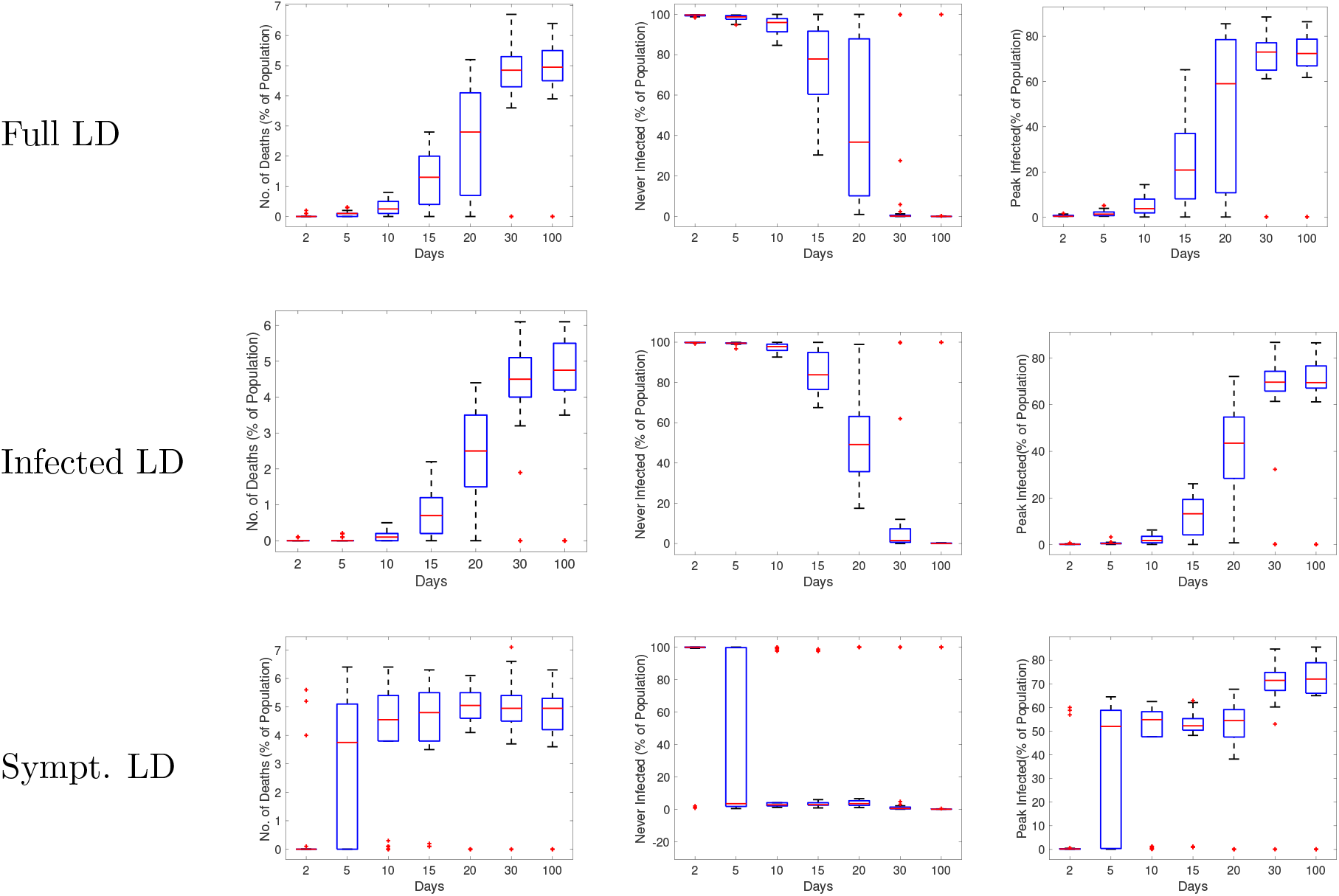
Statistical summaries of infection variables obtained using 30 runs of RAW-ALPS, plotted against the starting day of the full restrictions.

The first column shows the median percentage fatalities in the population for different lockdown types. The second column shows the fraction of uninfected population and the last column shows the peak infection rates. It can be seen that the results are very similar for a full lockdown and a lockdown of infected agents, but the lockdown of only the symptomatic agents is quite different. This last type of lockdown is found to be ineffective in containing the spread of the infection, except for when *T*_0_ = 2. These results suggest that a lockdown on infected agents can be an effective strategy in controlling the epidemic. Of course, given the asymptomatic nature of the disease, it is not possible to ascertain the infection status precisely. One can only estimate this status for a large population using accurate and extensive testing schemes.

In Fig. 15 we study the effect of changing *T*_1_ while *T*_0_ = 1 is kept fixed (and other experimental conditions being same as in the last experiment). The three panels show the fractions of deaths (left), the number of uninfected (middle), and the peak infection rates (right). Each curve in the panel corresponds to a different lockdown type: full LD or LD 1, infection LD or LD 2, and symptomatic LD or LD 3. The results show that for LD1 and LD2, a delay in lifting of the restrictions help control the epidemic quite effectively, while imposition of LD 3 is not as effective in the process. Interestingly, these results show that LD2 is more effective than LD 1. This maybe because in full lockdown the chances of susceptible agents co-inhabiting with the infected agents increase and this in turn increases the infection rates.

**Figure 15:**
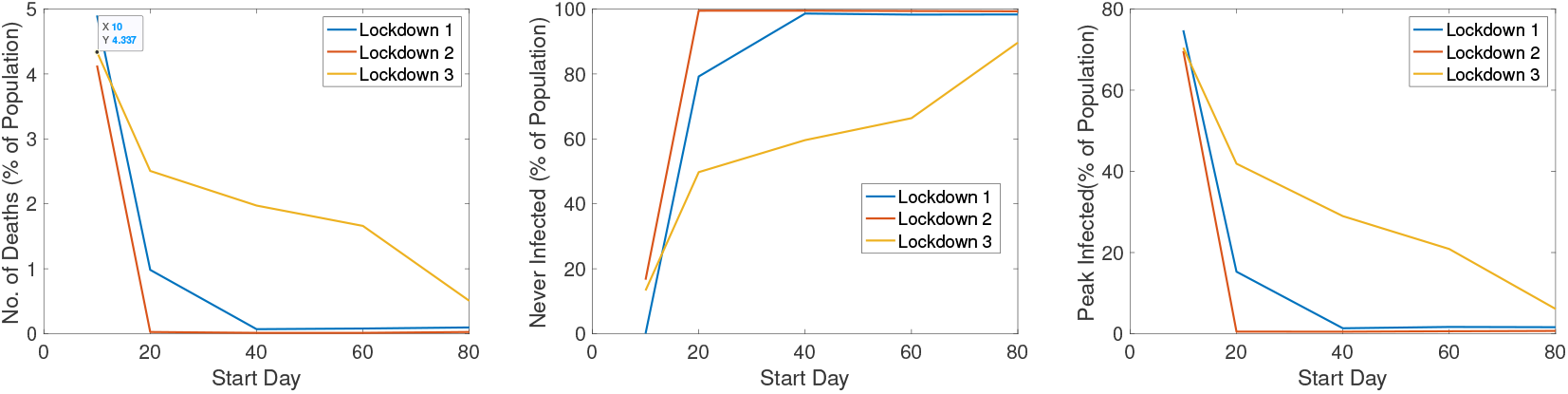
Statistical summaries of infection variables obtained using 30 runs of ALPS, plotted against *T*_1_, the reopening day.

## 5 Discussion & Conclusion

This paper develops an agent-based simulation model, called RAW-ALPS, for modeling spread of an infectious disease in a closed community. A number of simplifying and reasonable assumptions makes the RAW-ALPS efficient and effective for statistical analysis. The model is validated at a population level by comparing with the popular SIR model in epidemiology.

It concludes that a lockdown of only the infected agents is the most effective kind. However, this includes both symptomatic and asymptomatic agents with the later ones not easily detectable. This calls for regular and extensive testing of the population to isolate and restrict all infected agents, while allowing for free movements of all uninfected agents. Further, these results indicate that: (1) Early imposition of lockdown measures (right after the first infection) significantly reduce infection rates and fatalities; (2) Lifting of lockdown measures recommences the spread of the disease and the infections eventually reach the same level as the unrestricted community; (3) In absence of any extraneous solutions (a medical treatment/cure, a weakening mutation of the virus, or a natural development of agent immunity), the only viable option for preventing large infections is the judicious use of lockdown measures.

The strengths and limitations of RAW-ALPS model are the following. It provides an efficient yet comprehensive modeling of the spread of infections in a self-contained community, using simple model assumptions. The model can prove very useful in evaluating costs and effects of imposing different types of social lockdown measures in a society. In the current version, the initial placement of agents is set to be normally distributed with means given by their home units and fixed variance. This variance is kept large to allow for near arbitrary placements of agents in the community. In practice, however, agents typically follow semi-rigid daily schedules of being at work, performing chores, or being at home. Thus, at the time of imposition of a lockdown, the agents can be better placed in the scenes according to their regular schedules rather than being placed arbitrarily.

In terms of future directions, there are many ways to develop this simulation model to capture more realistic scenarios: (1) It is possible to model multiple, interactive communities instead of a single isolated community. (2) One can include typical daily schedules for agents in the simulations. A typical agent may leave home in the morning, spent time in the office during the day, and return to home in the evening. (3) It is possible to provide an age demographics to the community and assign immunity to agents according to their demographic labels [7]. (4) As more data becomes available in the future, one can change immunity levels of agents over time according to the spread and seasons. (5) In practice, when an agent is infected, he/she goes through different stages of the disease, associated with varying degrees of mobility [21]. One can introduce an additional variable to track these stages of infections in the model and change agent mobility accordingly.

## Data Availability

This manuscript contains simulated data and no real data is used in this paper.

## Acknowledgements

This research was supported in part by the grants NSF DMS-1621787 and NSF DMS-1953087.

**Listing of ALPS parameters**: Table 2 provides a listing of all the parameters one can adjust in ALPS to achieve different scenarios. It also provides some typical values used in the experiments presented in the paper.

---

**Table 1:**
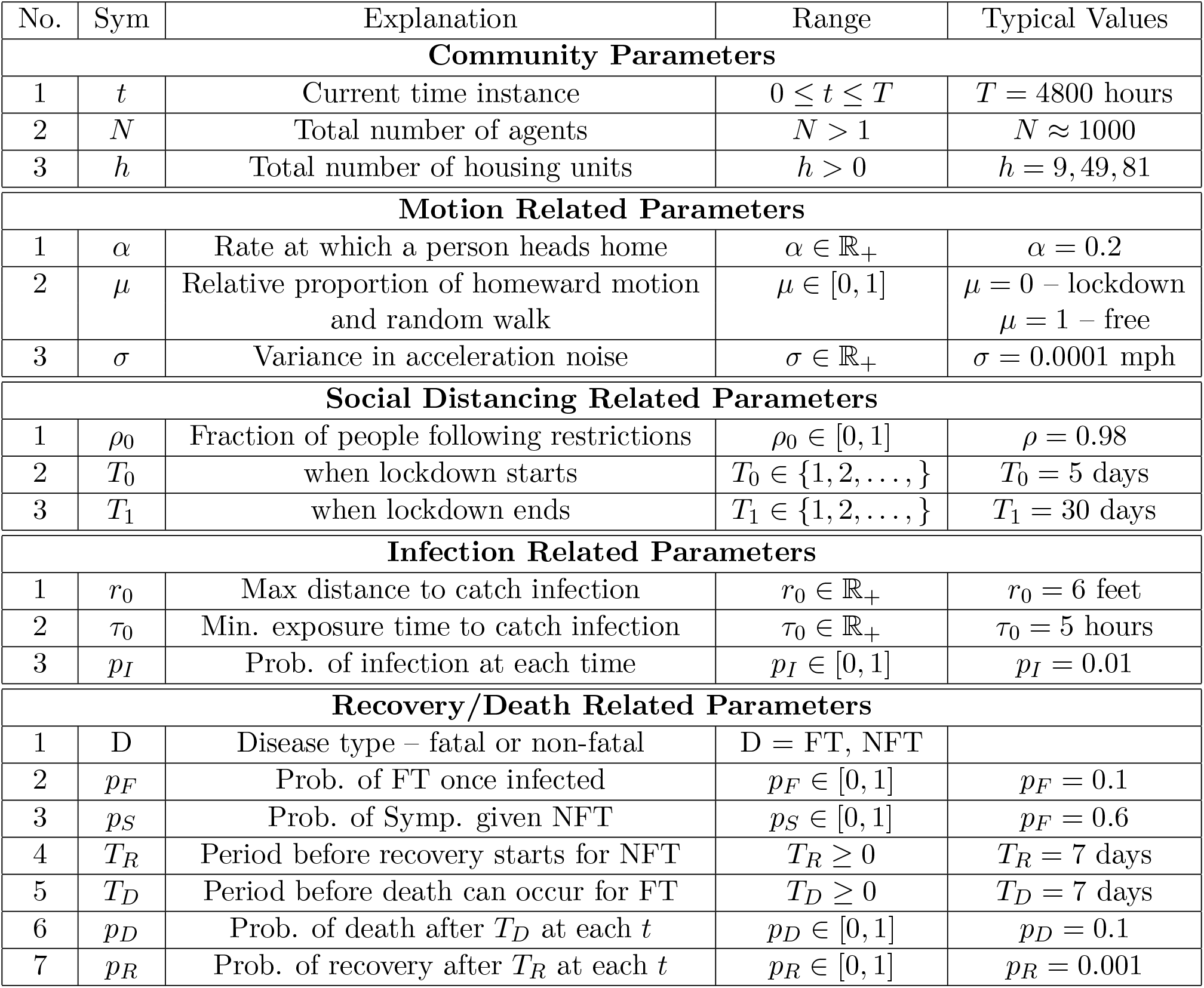
Listing of parameters associated with different model components of ALPS.

| No. | Sym | Explanation | Range | Typical Values |
| --- | --- | --- | --- | --- |
| **Community Parameters** |  |  |  |  |
| 1 | $t$ | Current time instance | $0 \leq t \leq T$ | $T = 4800$ hours |
| 2 | $N$ | Total number of agents | $N > 1$ | $N \approx 1000$ |
| 3 | $h$ | Total number of housing units | $h > 0$ | $h = 9, 49, 81$ |
| **Motion Related Parameters** |  |  |  |  |
| 1 | $\alpha$ | Rate at which a person heads home | $\alpha \in \mathbb{R}_+$ | $\alpha = 0.2$ |
| 2 | $\mu$ | Relative proportion of homeward motion and random walk | $\mu \in [0, 1]$ | $\mu = 0$ – lockdown<br>$\mu = 1$ – free |
| 3 | $\sigma$ | Variance in acceleration noise | $\sigma \in \mathbb{R}_+$ | $\sigma = 0.0001$ mph |
| **Social Distancing Related Parameters** |  |  |  |  |
| 1 | $\rho_0$ | Fraction of people following restrictions | $\rho_0 \in [0, 1]$ | $\rho = 0.98$ |
| 2 | $T_0$ | when lockdown starts | $T_0 \in \{1, 2, \dots, \}$ | $T_0 = 5$ days |
| 3 | $T_1$ | when lockdown ends | $T_1 \in \{1, 2, \dots, \}$ | $T_1 = 30$ days |
| **Infection Related Parameters** |  |  |  |  |
| 1 | $r_0$ | Max distance to catch infection | $r_0 \in \mathbb{R}_+$ | $r_0 = 6$ feet |
| 2 | $\tau_0$ | Min. exposure time to catch infection | $\tau_0 \in \mathbb{R}_+$ | $\tau_0 = 5$ hours |
| 3 | $p_I$ | Prob. of infection at each time | $p_I \in [0, 1]$ | $p_I = 0.01$ |
| **Recovery/Death Related Parameters** |  |  |  |  |
| 1 | D | Disease type – fatal or non-fatal | D = FT, NFT |  |
| 2 | $p_F$ | Prob. of FT once infected | $p_F \in [0, 1]$ | $p_F = 0.1$ |
| 3 | $p_S$ | Prob. of Symp. given NFT | $p_S \in [0, 1]$ | $p_F = 0.6$ |
| 4 | $T_R$ | Period before recovery starts for NFT | $T_R \geq 0$ | $T_R = 7$ days |
| 5 | $T_D$ | Period before death can occur for FT | $T_D \geq 0$ | $T_D = 7$ days |
| 6 | $p_D$ | Prob. of death after $T_D$ at each $t$ | $p_D \in [0, 1]$ | $p_D = 0.1$ |
| 7 | $p_R$ | Prob. of recovery after $T_R$ at each $t$ | $p_R \in [0, 1]$ | $p_R = 0.001$ |

